# Effective Aerosol Inoculation of Dose-Escalated Seasonal Influenza H3N2 Virus in Controlled Human Infection Model

**DOI:** 10.1101/2025.07.23.25332064

**Authors:** Nadine Rouphael, Ralph Tanios, Jessica Traenkner, Matthew D. Pauly, Nishit Shetty, Meredith J. Shephard, AJ Campbell, Christelle Radi, Shamika Danzy, Jin Pan, Flu CHIM Study Group, Andrew P. Catchpole, Alex Mann, Joshua F. Detelich, Anice C. Lowen, Linsey C. Marr, Seema S. Lakdawala

## Abstract

Human challenge models (CHIMs) are instrumental in advancing influenza research but have traditionally relied on intranasal inoculation, which does not mimic the natural aerosol transmission of the virus. We conducted a dose-escalation influenza CHIM study to evaluate the safety and feasibility of two modern aerosol delivery systems: a flow-focusing monodisperse aerosol generator (FMAG) and a medical nebulizer. Fourteen healthy adults aged 18–49 years were exposed to influenza A/Perth/16/2009 (H3N2) in a controlled inpatient setting. Infection rates were 75% (3/4) with FMAG and 50% (2/4) with the nebulizer at the higher dose. Infections were self-limited, with sinus congestion, rhinorrhea, and cough being the most common symptoms. No serious adverse events occurred. Viral shedding was reproducible across respiratory sites, and seroconversion occurred in 33% of infected participants. Symptom timing and viral kinetics were comparable to those observed in prior intranasal CHIMs. Participants receiving nebulizer-delivered virus showed earlier viral detection in the oral cavity, suggesting broader airway deposition. These findings demonstrate that aerosol influenza challenge is both safe and effective and can simulate natural infection more accurately than intranasal delivery. This reintroduction of aerosolized influenza challenge provides a robust platform for studying transmission dynamics, tissue-specific immune responses, and for evaluating next-generation vaccines and therapeutics under conditions that better approximate real-world exposure.

**One Sentence Summary:** We demonstrated controlled human infection with dose-escalated H3N2 influenza virus via two safe and effective aerosolization routes that provide more accurate simulation of real-world exposure than intranasal delivery for studying influenza transmission, tissue-specific immune responses, and vaccines/therapeutics.

## INTRODUCTION

Controlled Human Infection Models (CHIMs) have been essential for advancing our understanding of influenza pathogenesis, immune responses, and vaccine efficacy *(1)*. To ensure safety and consistency, intranasal inoculation became the standard, though it circumvents the natural aerosolized route of transmission, limiting its ability to fully model community acquired infection.

Inoculation with aerosolized influenza virus was first reported in 1937 *(2)*, and this approach was instrumental in shaping early understanding of influenza immunity and vaccine responses *(3-5)*. Subsequent studies demonstrated that inhaled aerosols could initiate infection with much lower doses than required via intranasal inoculation *(6)*, and that the site of deposition influenced disease manifestation *(7)*. Indeed, a comparative analysis suggested that aerosol challenges may be up to 20 times more efficient at inducing infection than intranasal inoculation, likely due to deeper respiratory tract deposition *(8)*. Although mid-20th-century aerosol challenge studies reported high attack rates—up to 81% *(7)*—concerns over safety and variability *(6)* led to the method’s decline *(9,10)*. Nevertheless, aerosol inoculation more closely mimics natural infection, allowing deposition of virus-laden aerosol particles in both the upper and lower respiratory tracts *(11-14)*. With advances in aerosol delivery systems and stricter safety protocols, there is renewed interest in aerosol-based human challenge models to better simulate natural transmission, pathogenesis, and immune responses.

Here, we conducted a dose-escalation study using two modern aerosol delivery systems to evaluate the safety and feasibility of influenza virus challenge via inhalation. Our primary aim was to re-establish an aerosol-based CHIM that more closely mirrors natural respiratory transmission, enabling mechanistic studies of viral shedding, early mucosal infection, and host response. This model provides a foundation for exploring factors that influence transmission and for testing interventions designed to interrupt the spread of respiratory pathogens.

## RESULTS

### Participant demographics and exposure doses

Fourteen healthy adults (ages 21–49 years, 71% female and 50% identifying as non-White) were enrolled and inoculated with influenza A/Perth/16/2009 (H3N2) virus via aerosol using either the FMAG or a medical nebulizer (Fig. S1 and Table S1). Estimated exposure doses were derived from spirometry and aerosol sampling measurements (Table).

### Infection rates

Infection rates varied by dose and device. At the lower dose (3.6 × 10^3^ TCID_50_/mL), FMAG inoculation resulted in 0/2 infections, while nebulizer delivery led to infection in 1/4 participants (25%). At the higher dose (3.6 × 10^4^ TCID_50_/mL), FMAG produced infections in 3/4 participants (75%) and the nebulizer in 2/4 (50%).

### Symptom scores

Symptoms were generally mild and self-limited. FMAG-associated MMID cases most commonly reported runny nose, sinus congestion, and headache, whereas nebulizer-associated MMID cases had cough, fatigue, and decreased appetite, the latter included in the “headache/body aches/fatigue” symptom category in Fig. 1. Only one participant (F097) developed a fever of 38.0°C. Symptom severity tracked with viral load in most participants. Participant F107 reported the highest symptom burden, primarily cough, headache, sore throat, and fatigue, which preceded PCR positivity and continued after oseltamivir initiation on Day 5. Despite higher symptom scores, viral loads in this participant were comparable to others in the nebulizer cohort.

**Fig. 1.**
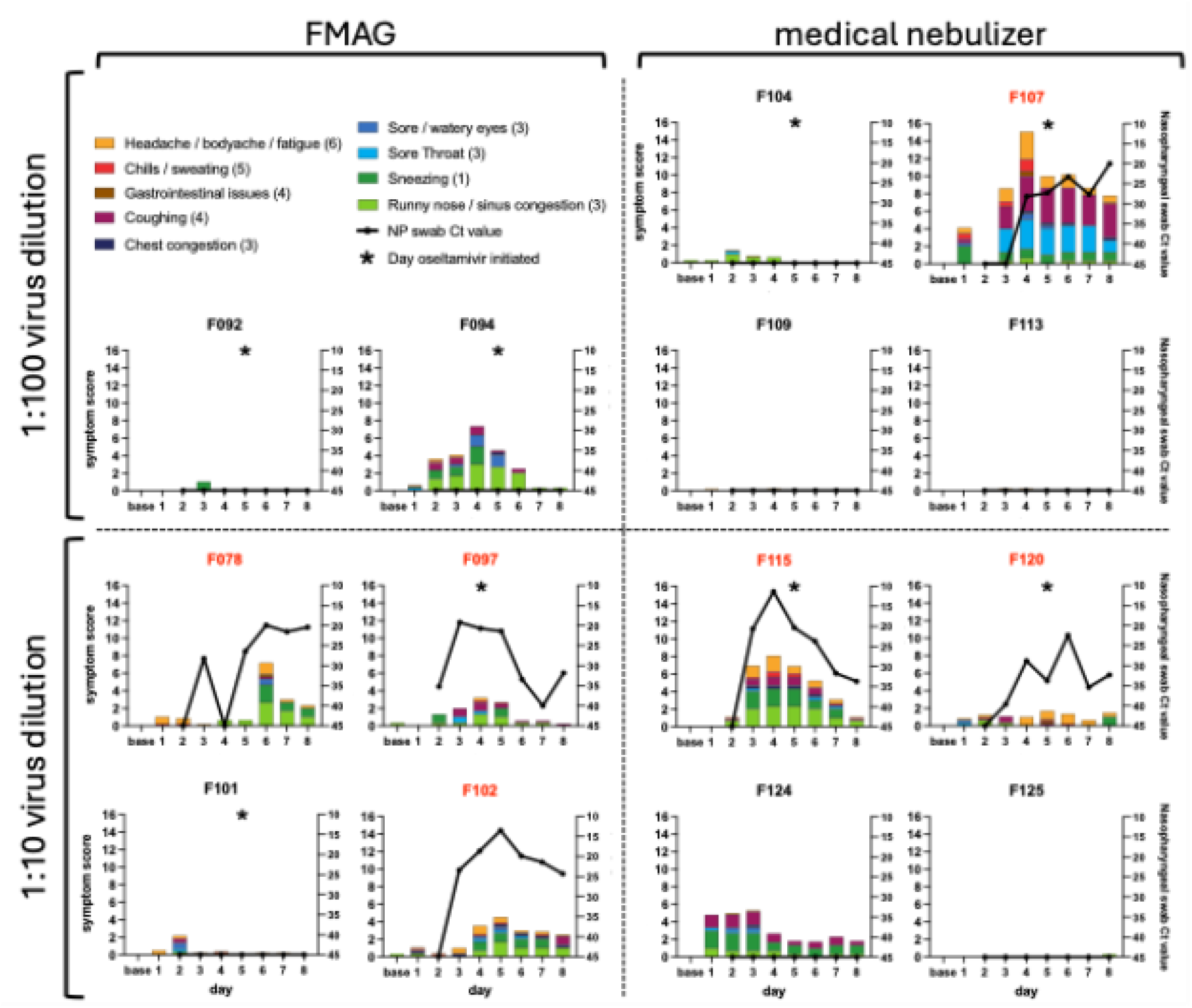
Symptom scores and nasopharyngeal swab titers. The mean daily symptom scores for nine stacked, color-coded symptom categories are plotted on the left axis. Scores for each symptom range from 0-4 and the number of symptoms included in each category is indicated in parentheses in the legend. Participants were inoculated on Day 1. Baseline symptoms were collected on Day 1 prior to inoculation. The cycle threshold (Ct) value obtained from daily testing of nasopharyngeal swab samples is plotted as a black line on the right axis. Samples in which influenza was not detected were given a Ct value of 45. Participants are grouped by aerosol generation device with the FMAG (flow-focusing monodisperse aerosol generator) on the left and the medical nebulizer on the right, and by virus dilution: 1:100 at the top and 1:10 at the bottom. Red participant identifiers indicate participants who met the MMID case definition. Asterisks denote the first day that daily oseltamivir phosphate doses were administered to a participant.

### Viral titers

Viral RNA was detected in multiple upper airway sites, often persisting throughout the 8-day inpatient period (Fig. 2). Influenza virus was first detected on Day 2 for participant F097 and Day 3 for the other five participants with MMID. Despite low levels of influenza genetic material in samples from participants F113 and F124, these detections did not occur in nasal samples and, therefore, these participants did not meet the criteria for MMID classification. In participant F107, infectious virus was detectable in saliva and oropharyngeal samples prior to nasopharyngeal detection, aligning with symptom onset. Across MMID cases, viral loads peaked between Days 2–3 post-inoculation (Fig. 3). Overall, viral replication in nasopharyngeal swab samples after aerosol inoculation was similar to or slightly delayed when compared to inoculation via the intranasal route^15^ (Fig. 4A). Symptom timing and overall severity were generally similar to those observed following intranasal inoculation, although in some cases, symptom onset appeared slightly delayed, consistent with the timing of viral load detection (Fig. 4B).

**Fig. 2.**
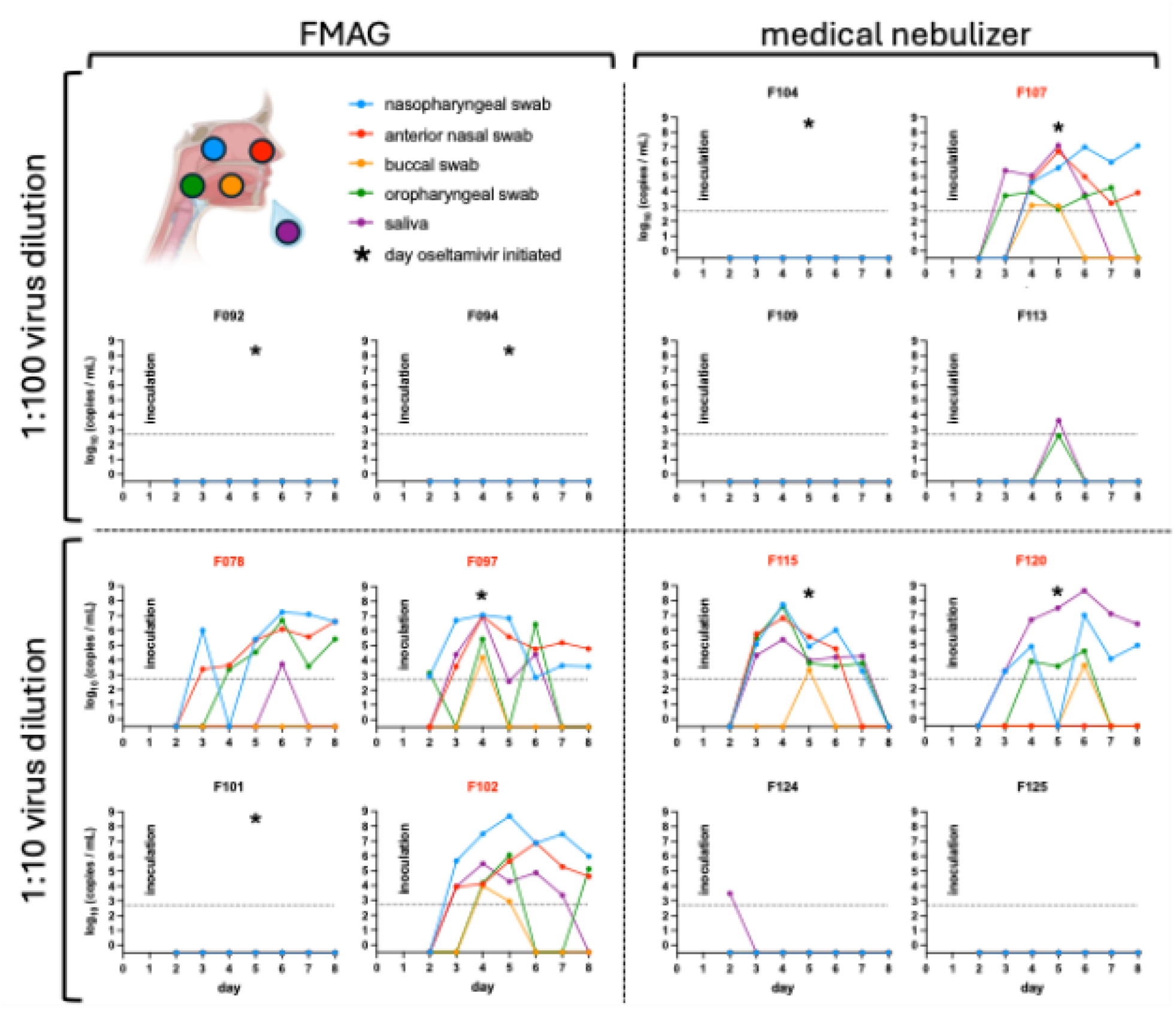
Quantification of influenza viral RNA at different anatomical sites over time. The amount of influenza viral RNA was determined by quantitative reverse-transcription PCR (qRT-PCR) of daily nasopharyngeal swab (blue), anterior nasal swab (red), buccal swab (yellow), oropharyngeal swab (green), and saliva (purple) samples for all participants. The limit of detection is indicated by a dotted line. Participants are grouped by aerosol generation device with the FMAG (flow-focusing monodisperse aerosol generator) on the left and the medical nebulizer on the right, and by virus dilution: 1:100 at the top and 1:10 at the bottom. Inoculations occurred on Day 1. Red participant identifiers indicate those who met the MMID case definition. Asterisks denote the first day that daily oseltamivir phosphate doses were administered to a participant.

**Fig. 3.**
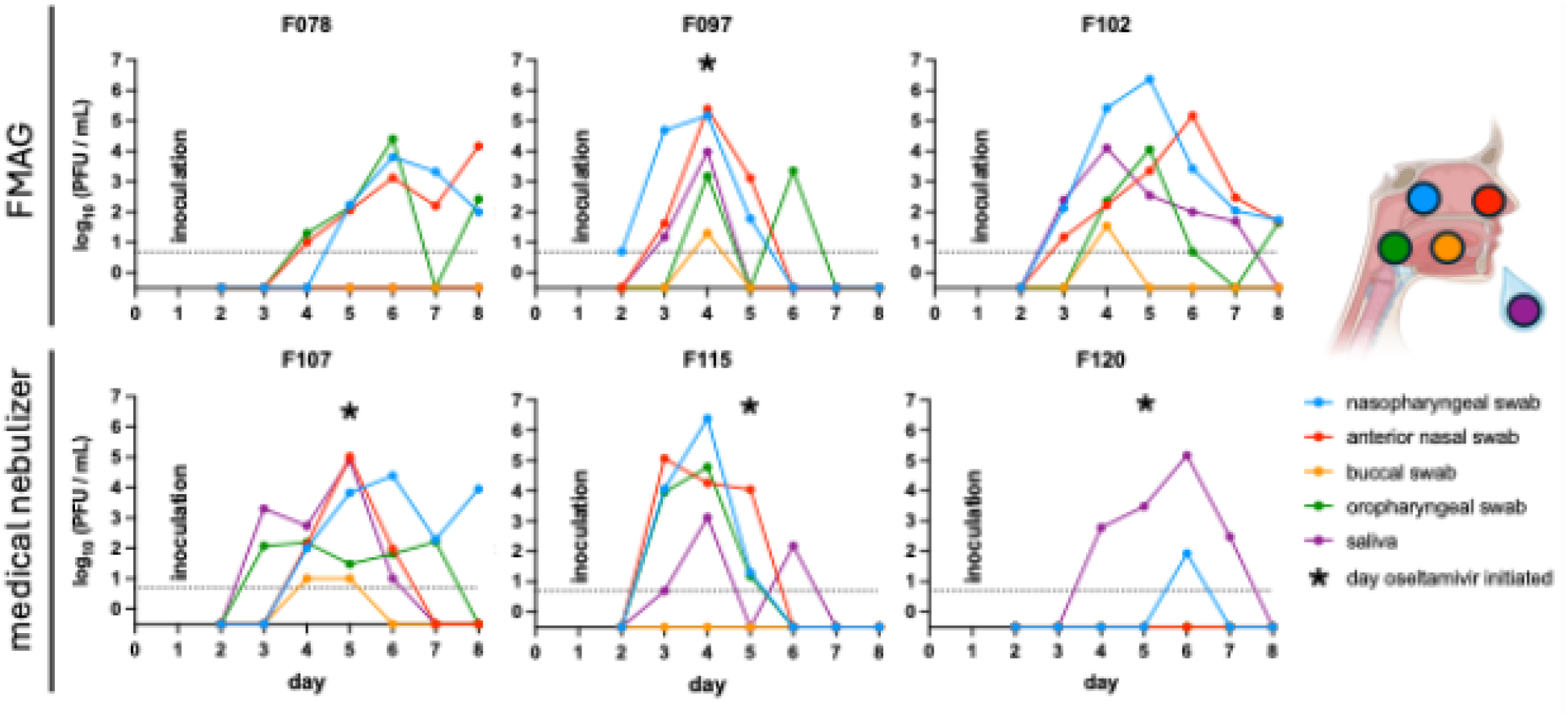
Infectious virus titers at different anatomical sites. The amount of infectious virus was determined by plaque assay of daily nasopharyngeal swab (blue), anterior nasal swab (red), buccal swab (yellow), oropharyngeal swab (green), and saliva (purple) samples for the six participants with MMID. The limit of detection was 5 PFU/mL and is indicated by a dotted line. Participants are grouped by aerosol generation device with the FMAG (flow-focusing monodisperse aerosol generator) on the top and the medical nebulizer on the bottom. Inoculations occurred on Day 1. Asterisks denote the first day that daily oseltamivir phosphate doses were administered to a participant.

**Fig. 4.**
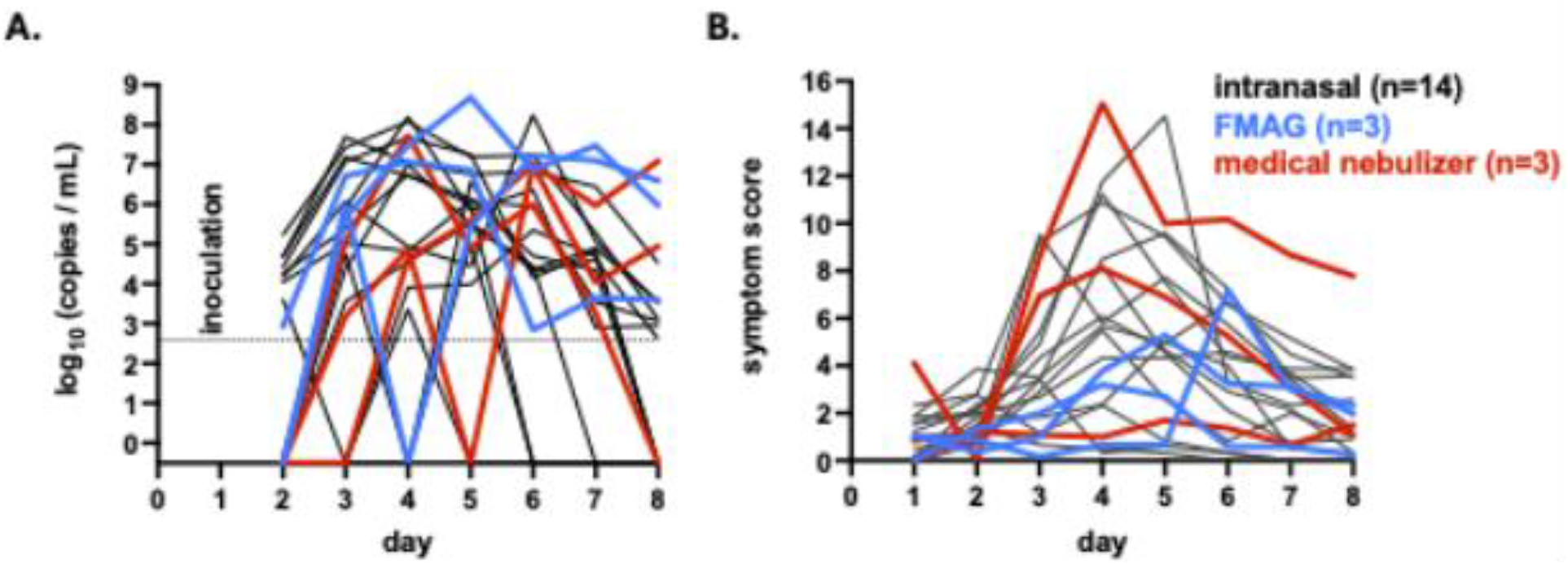
Comparison of viral replication kinetics and symptoms between intranasal and aerosol inoculation routes. **(A)** The influenza genome copies measured in nasopharyngeal swab samples tested by quantitative reverse-transcription PCR (qRT-PCR) assay. Participants were inoculated via either the intranasal route^15^ (black, n=14, including six from reference 15 and nine unpublished) or the aerosol route using an FMAG (flow-focusing monodisperse aerosol generator) (blue) or medical nebulizer (red) on study Day 1. Only participants with MMID are shown. The assay’s limit of detection is indicated by the dashed line. **(B)** The sum of the mean FLU-PRO symptom responses for the nine symptom categories listed in Figure 1, each scored on a scale from 0 to 4, for the same participants as in panel A. Responses for study Day 1 were collected in the evening, after participants were inoculated in the morning.

### Seroconversion rates

Seroconversion was limited. Among FMAG participants, 1/6 (17%) had ≥4-fold HAI titer increases by Day 31; in those with MMID, 1/3 (33%) seroconverted. Among nebulizer participants, 1/8 (13%) seroconverted overall, and 1/3 (33%) with MMID (Fig. S2). These muted systemic responses occurred despite robust viral shedding.

### Safety outcomes

Safety outcomes were favorable. Two related AEs occurred in the FMAG group (grade 1 fever, which resolved the same day, and a grade 2 lymphopenia which resolved in 11 days). The nebulizer group had six related AEs, including grade 3 lymphopenia in one participant, all of which resolved. No serious adverse events, influenza-related complications, or abnormalities in ECG, PFT, or chest imaging were observed.

## DISCUSSION

This study reintroduces human challenge with aerosolized influenza A virus and demonstrates that two modern aerosol delivery systems—FMAG and medical nebulizer—can safely induce mild, self-limited infection and reproducible viral shedding in a controlled setting. Using a dose-escalation approach, we found infection rates of 50–75% at higher viral loads, with no serious adverse events and symptom severity comparable to traditional intranasal CHIMs. These results establish feasibility for using aerosol challenge models to better simulate natural transmission and explore respiratory mucosal immunity, shedding dynamics, and the early host response.

Early influenza challenge models explored aerosolized viral inoculation. Notable examples include a study published in 1946 where atomization of allantoic fluid containing greater than 10^7^ chick embryo ID_50_ of the PR-8, F-12, or F-99 influenza A strains yielded attack rates between 62.5 and 81.8% *(6)*. Another example, from 1966, showed that 7 of 14 subjects (50%) with neutralizing antibody titers ≤1:40, exposed to between 1 and 59 inhaled TCID_50_ of A/Bethesda/10/63 (H2N2) exhibited illness, shed virus, and/or seroconverted *(6)*. The doses in our study fall between those administered in these historical reports. Inoculations using a 3·6×10^3^ TCID_50_/mL inoculum solution, exposing participants to between 20 and 1000 PFU (plaque-forming units) led to infection in 1 of 6 (16.7%) participants. A ten-fold more concentrated inoculum exposed participants to between 1000 and 30,000 PFU, resulting in infection in 5 of 8 participants (62.5%). Fever was the most commonly assessed and reported symptom in prior studies. In Henle et al., 58 of 65 (89%) individuals who inhaled influenza challenge virus experienced fever, with a maximum reported temperature of 40°C (104°F) *(7)*. In Alford et al., the mean maximum temperature among the four symptomatic participants was 38·8°C (101·8°F) despite frequent aspirin administration *(6)*. In contrast, only 1 of the 6 participants with MMID (16.7%) in our study exhibited fever. This difference may be attributable to strain variation or improved supportive care. Other notable symptoms in these studies included headache, malaise, myalgia, nasal obstruction, rhinorrhea, sore throat, cough and, less commonly, flushing of the face and redness of the anterior pillars of the soft palate. In rare instances, more serious symptoms were noted in these previous studies using aerosol delivery of influenza, including multiple bouts of emesis lasting 24 hours in one subject and inspiratory and expiratory wheezes lasting six days in another *(7)*. In contrast, our study identified only mild symptoms associated with aerosol inoculation of influenza. However, differences in virus strains, inoculum dose, aerosolization devices, and participant baseline immunity may account for variations in illness severity between past and current studies *(6,7)*. Our study builds upon these historical findings by implementing rigorous safety measures, controlled dose escalation, and optimized aerosolization delivery systems, ensuring participant safety while maximizing model reproducibility.

We selected the FMAG device for our initial studies due to its ability to generate particles in the 8-11 μm size range that would primarily deposit in the upper respiratory tract; less than 10% of them would deposit deeply in the lungs (i.e, the alveoli). In contrast, the particles produced by the medical nebulizer are primarily smaller than 5 µm and therefore expected to penetrate throughout the lower respiratory tract in addition to depositing in the upper respiratory airways. The utility of these two approaches may differ across settings: the FMAG is more complex to operate and less readily available compared to a standard medical nebulizer. Despite these differences, inoculations using both aerosolization methods induced relatively mild disease in the exposed participants and shedding of infectious virus in multiple respiratory sites. Of note, in the three participants infected via the medical nebulizer, viral load was detectable in the oral cavity prior to or on the same day as detection in nasopharyngeal swabs suggesting that infection using this approach was initiated, at least in part, outside the nasal cavity. Interestingly, despite robust viral infection, few participants seroconverted, suggesting a more complex relationship between systemic antibody induction and seasonal influenza infection.

There are several limitations to this study. First, the small sample size limits generalizability and precluded stratified analyses by sex, gender, race, or ethnicity. Second, we evaluated only two aerosolization delivery methods and a single virus strain. Findings may differ with other devices or strains. Third, while considering the utility of this modified CHIM approach for applications where symptomatology is a primary outcome, it is notable that the MMID observed in our study was milder than medically attended influenza cases and consistent with other CHIM studies using intranasal inoculation in a young, healthy population. Fourth, the dose estimation relied on sampling and analysis of aerosolized virus, which is known to be inefficient. Infectivity may be partially lost during such collection and processing, potentially leading to an underestimate of the actual dose of infectious virus delivered to participants. Finally, the inoculum was derived from an egg-adapted virus, which may preferentially bind to α2,3-linked sialic acids, associated with enhanced infectivity in the lower respiratory tract. This receptor-binding preference could influence outcomes when virus is delivered via aerosol, as opposed to intranasal inoculation, where α2,6-linked sialic acids dominate and support replication in the upper respiratory tract.

Future studies will examine how immunological responses differ by mode of viral delivery, comparing intranasal and aerosol challenge models to gain insight into the impact of viral distribution on tissue-resident and systemic innate and adaptive immunity. In particular, this platform may be useful for evaluating mucosal vaccine candidates, antivirals that aim to block transmission, and immunologic correlates of protection at the site of virus entry. By simulating the natural route of influenza transmission under tightly controlled conditions, aerosol challenge models offer a valuable translational tool for respiratory virus research, bridging basic virology with intervention development.

## MATERIALS AND METHODS

### Study Design

This study evaluated the safety and feasibility of aerosol challenge using influenza A/Perth/16/2009 (H3N2), a virus that has been previously administered intranasally to participants (*15,16*). In the interest of safety, inoculations were first performed using an aerosolization method (Flow Focusing Monodisperse Aerosol Generator (FMAG), TSI Incorporated, Shoreview, MN) that produced particles with an aerodynamic diameter of 8-11 μm. Particles of this size primarily deposit in the upper respiratory tract (*12,13*). Once safety was documented for inoculations with larger diameter aerosols, inoculations then proceeded using a medical nebulizer (Medline Industries, Northfield, IL and Philips Respironics, Murrysville, PA) that produced particles almost entirely <5 μm, with a number mode <1 μm. Particles of this size can deposit in both the upper and lower respiratory tracts. The virus was aerosolized into a fit test hood (3M, St. Paul, MN) worn by the participants for 10-20 minutes (see Supplement). For each aerosolization device, cohorts of two participants were exposed to a dose approximately equivalent to a 1:100 dilution of the standard intranasal dose of 3.6 × 10^5^ TCID_50_ (*15*). Due to dilution of aerosolized virus with background air in the fit test hood and the less-than 100% deposition efficiency of particles in the respiratory tract, the actual dose delivered to the participants was lower. Dose validation was performed to confirm these doses (see Supplement). Initial participants received abortive antiviral therapy (oseltamivir phosphate, 75 mg taken twice daily for 5 days) either 48 hours after a positive PCR test or by 4 days post challenge, or if clinical deterioration occurred. If mild to moderate influenza disease (MMID; for definition, see Supplement) occurred in 50% of participants, the same dose was then tested in a new cohort of two participants without early antiviral treatment. If MMID did not occur, a new cohort of two participants was exposed to a 10-fold higher dose and received antiviral therapy as indicated above. The study used predefined halting criteria (see Supplement). Safety data, including symptoms, lab results, chest radiograph imaging, electrocardiograms (ECGs), pulmonary function tests (PFTs), and adverse events, were reviewed post-cohort by an Independent Safety Monitor (ISM) to guide dose escalation (see Supplement).

### Participants

Healthy, non-pregnant adult participants (18–49 years old) were enrolled following pre-screening for hemagglutination inhibition (HAI) titers ≤1:40 and a comprehensive health evaluation (*15*) between February 2024 and March 2025 (ClinicalTrials.gov identifier NCT05332899). All participants provided informed consent and passed an assessment of understanding prior to study procedures. Participants were admitted to a designated inpatient human challenge unit with quarantine measures at Emory University Hospital for controlled exposure and monitored from study Days 0 to 8 and challenged on Day 1. Participants self-reported sex assigned at birth, gender identity, race, and ethnicity during pre-screening as part of standard demographic data collection. Categories were based on U.S. Office of Management and Budget standards, allowing multiple selections. These data were used for descriptive purposes and not for subgroup analysis, given the small sample size. The study was approved by the Emory IRB (protocol STUDY00000083) and the Food and Drug Administration (FDA IND #19579).

### Virus Strain

The study used influenza A/Perth/16/2009 (H3N2) virus, produced under Good Manufacturing Practice (GMP) standards and manufactured by Meridian Life Sciences (Memphis, TN) on behalf of hVIVO (*16-18*). The virus was stored at −70°C to −80°C in single-use vials. The virus is known to be susceptible to neuraminidase inhibitors (*19*).

### Monitoring and Sampling

Participants underwent frequent monitoring in the hospital for clinical symptoms, vital signs, and viral shedding. Symptom severity was recorded daily using the InFLUenza Patient-Reported Outcome (FLU-PRO) diary (*15*) from study Days 1–15. Vital signs were measured three times daily, accompanied by targeted daily clinical exams. Safety assessments included complete blood count (CBC) with differential, creatinine, alanine aminotransferase (ALT), and urinalysis on Days 2 and 4; chest radiography and PFTs on Day 7; and ECGs on Days 3 and 6. Adverse events (AE) were assessed from the day of challenge until Day 15 for solicited AE, until approximately Day 31 for unsolicited AE, and until Day 91 for serious AE, using Common Terminology Criteria for Adverse Events (CTCAE) Version 5.0 (November 27, 2017). Daily nasopharyngeal swabs were collected during quarantine for PCR-based viral detection (see Supplement). Assessment of both infectious and molecular viral loads at multiple upper respiratory sites was also conducted daily (see Supplement). Antibody measurements by HAI assay^15^ were conducted on serum samples collected on study Days 1, 15 and 31.

## Data Availability

All data in the main text or the supplementary materials and available online in a figshare repository at https://figshare.com/s/e9faf12d099b4b2d8bc9; private link

## ACKNOWLEDGEMENTS

We would like to thank the participants who enrolled in our study and Emory University Hospital Nursing staff for their technical assistance. We thank members of the Hope Clinic, Lakdawala Lab, Lowen Lab, and Marr lab for useful discussions.

## AUTHOR CONTRIBUTIONS

Conceptualization: NR, MDP, ACL, LCM, SL

Methodology: NR, NS, JP, ACL, LCM

Investigation: NR, RT, JT, MDP, MJS, CR, SD, GQ, KP, NVM, MV, KBu, KBr, MG, HN, AL, VS, CZ

Formal analysis: MDP, MJS, AC, SD, GQ, KP, NVM, MV, KBu, KBr, MG, HN, AL

Validation: MDP, NS, AC, SL

Visualization: NR, RT, NS, CR, JP, ACL

Funding acquisition: NR, SL

Project administration: NR, JT

Supervision: NR, SL

Resources: AC, AM, JFD, LCM, CSK, AKM, DAG, MFE, ECF

Writing – original draft: NR, ACL, LCM, SL

Writing – review & editing: NR, RT, JT, MDP, NS, MJS, AC, CR, SD, JP, AC, AM, JFD, ACL, LCM, SL, GQ, KP, NVM, MV, KBu, KBr, MG, HN, AL, VS, CZ, CSK, AKM, DAG, MFE, ECF

## Data and materials availability

All data are available in the main text or the supplementary materials and available in a figshare repository (https://figshare.com/s/e9faf12d099b4b2d8bc9; private link).

## Flu CHIM Study Group

Department of Pathology and Laboratory Medicine and the Department of Medicine, Division of Infectious Diseases, at Emory University School of Medicine in Atlanta, Georgia, USA: Colleen S Kraft

Division of Infectious Diseases, at Emory University School of Medicine in Atlanta, Georgia, USA: Aneesh K Mehta

Georgia Clinical and Translational Science Alliance, Emory University School of Medicine, Atlanta, GA, USA: Dalia A Gulick

Division of Cardiology, Department of Medicine, Emory University School of Medicine, Atlanta, GA, USA: Mikhael F El-Chami

Department of Pathology and Laboratory Medicine, Emory University School of Medicine, Atlanta, GA, USA: Eric C Fitts

Hope Clinic, Division of Infectious Diseases, Department of Medicine, Emory University, Decatur, Georgia, USA: Veronica Smith and Cecilia Zhang

Department of Microbiology and Immunology, Emory University School of Medicine, Atlanta, Georgia, USA: Grace Quirk, Kayle Patatanian, Nahara Vargas-Maldonado, Michelle Vu, Kayla Brizuela, Kaitlyn Bushfield, Matthew Gaddy, Hanh D Nguyen

Department of Civil and Environmental Engineering, Virginia Tech, Blacksburg, VA, USA: Alexandra Longest

## FIGURES AND TABLES

**Table 1.** Participant outcomes by aerosolization device and exposure. PTID=participant ID; Ct= cycle threshold, MMID= mild to moderate influenza disease, PCR= polymerase chain reaction; PFU = plaque-forming units, TCID_50_ =50% tissue culture infectious dose; HAI= hemagglutination inhibition; FMAG= flow-focusing monodisperse aerosol generator; Abnormal safety testing includes any grade 1 or above for complete blood count (CBC) with differential, creatinine, alanine aminotransferase (ALT) or a decrease in FEV_1_ (Forced Expiratory Volume in one second) of 10% or more from pre-challenge to post-challenge measurements.

| Aerosolization Device | Virus inoculum dilution (TCID<sub>50</sub>/mL) | PTID | Estimated Exposure (PFU) | Symptoms | Presence of MMID (Lowest PCR Ct value) | Abnormal Safety Testing | 4-fold rise in HAI titers |
| --- | --- | --- | --- | --- | --- | --- | --- |
| **FMAG** | 3·6 x 10<sup>3</sup> | F092 | 2x10<sup>1</sup> | - | No |  | No |
|  |  | F094 | 2x10<sup>1</sup> | - | No |  | No |
|  | 3·6 x 10<sup>4</sup> | F097 | 1x10<sup>3</sup> | Congestion<br>Productive Cough | Yes (19·2) | Grade 2 decrease in lymphocyte count | Yes |
|  |  | F101 | 6x10<sup>3</sup> | - | No |  | No |
|  |  | F078 | 9x10<sup>3</sup> | Rhinorrhea<br>Congestion<br>Headache<br>Fatigue | Yes (19·9) |  | No |
|  |  | F102 | 3x10<sup>4</sup> | Rhinorrhea<br>Congestion<br>Sinus Pressure<br>Headache<br>Productive Cough | Yes (13·5) |  | No |
| **Medical Nebulizer** | 3·6 x 10<sup>3</sup> | F104 | 1x10<sup>3</sup> | - | No |  | No |
|  |  | F107 | 6x10<sup>2</sup> | Headache<br>Decreased appetite<br>Sneezing<br>Cough | Yes (20) | Grade 3 decrease in lymphocyte count | No |
|  |  | F109 | 5x10<sup>2</sup> | - | No |  | No |
|  |  | F113 | 9x10<sup>2</sup> | - | No |  | No |
|  | 3·6 x 10<sup>4</sup> | F115 | 9x10<sup>3</sup> | Cough<br>Sinus Pressure<br>Decreased appetite<br>Fatigue<br>Rhinorrhea<br>Nasal Congestion | Yes (11·3) | Grade 2 decrease in lymphocyte count | Yes |
|  |  | F120 | 1x10<sup>4</sup> | Decreased appetite<br>Fatigue<br>Cough<br>Nasal Congestion | Yes (22·4) |  | No |
|  |  | F124 | 6x10<sup>3</sup> | - | No |  | No |
|  |  | F125 | 4x10<sup>3</sup> | - | No |  | No |

