## Supplementary Materials for "Effective Aerosol Inoculation of Dose-Escalated Seasonal Influenza H3N2 Virus in Controlled Human Infection Model"

**LIST OF SUPPLEMENTARY MATERIALS**

Materials and Methods

Fig. S1 to S3

Table S1

### SUPPLEMENTARY MATERIALS

#### Material and Methods

**Mild to Moderate Influenza Disease (MMID)** is defined as:

- Influenza A virus shedding detected by PCR from a nasal swab, plus
- One or more symptoms related to the study agent, including:
    - Arthralgia
    - Chills
    - Conjunctivitis
    - Nasal/sinus congestion
    - Decreased appetite
    - Diarrhea
    - Dry cough
    - Dyspnea/shortness of breath
    - Fatigue/tiredness
    - Fever (>38·0°C)
    - Headache
    - Lymphopenia (<1000 cells/mL)
    - Myalgia
    - Nausea
    - Oxygen saturation decrease by ≥3% from baseline and ≤92%
    - Cough
    - Nasal discharge
    - Sore throat
    - Sweats

**Study halting rules**

- Any participant experiences laryngospasm, bronchospasm, or anaphylaxis within one day after challenge.
- Two or more participants experience the same related grade 3 adverse event of any kind for more than 72 hours after challenge through quarantine period.
- Any participant experiences generalized urticaria (defined as occurring at more than two body parts) within 3 days after challenge and related to challenge.
- Any participant experiences a related serious adverse event.

- Any participant develops influenza complications during quarantine.
- Any other reason as deemed by the Principal Investigator.

**Individual stopping rules**

- Participant becomes noncompliant.
- Medical disease or condition, or new clinical finding(s) for which continued
  - participation, in the opinion of the investigator might compromise the safety of the
  - participant, interfere with the participant's successful completion of this study, or interfere with the evaluation of responses.
- Participant lost to follow-up.
- Participant becomes pregnant.

**Dose Escalation Criteria**

- Prior to any dose escalations in subsequent cohorts, the aggregate safety data through Day 15 post inhalation challenge is reviewed for all enrolled study participants of the current cohort. The review is conducted by the designated ISM and includes a review of lab results, imaging results, ECG's, PFT's, solicited and unsolicited AEs, and SAEs. If there are no significant safety concerns, dose escalation in the next cohort may proceed.
- Consultation with regulatory authorities may occur if indicated by any review. The ISM may recommend evaluation of additional participants at the current dose level and reassessment for safety before proceeding to a higher dose level if warranted by the interim safety review. Once influenza positivity is observed in either participant in the absence of abortive antiviral therapy and any significant safety events, the dose is set and no further escalation occurs.

**Symptom scores**

Responses to the 32 symptom types on the FLU-PRO questionnaire were converted to a 0–4 scale, with 0 corresponding to "not at all," "never," or "0 times," and 4 corresponding to "very much," "always," or "4 or more times." Symptoms were binned into nine categories, and mean values were calculated daily.

These categories include:

- **Runny nose/sinus congestion** (runny or dripping nose, congested or stuffy nose, and sinus pressure)
- **Sneezing**
- **Sore throat** (scratchy or itchy throat, sore or painful throat, and difficulty swallowing)
- **Sore/watery eyes** (teary or watery eyes, sore or painful eyes, and eyes sensitive to light)
- **Chest congestion** (trouble breathing, chest tightness, and chest congestion)
- **Coughing** (dry or hacking cough, wet or loose cough, coughing, and coughed up mucus or phlegm)
- **Gastrointestinal issues** (felt nauseous, stomach ache, number of vomiting episodes, and number of diarrhea episodes)
- **Headache/body ache/fatigue** (head congestion, headache, sleeping more than usual, weak or tired, body aches or pains, and decreased appetite)
- **Chills/sweating** (chills or shivering, felt cold, felt hot, sweating, and felt dizzy)

**Sample Collection**

Nasopharyngeal, anterior nasal, buccal, and oropharyngeal samples were collected using swabs placed into transport tubes (BD Universal Viral Transport Collection Kit, Cat. No. 220526, Fisher Scientific, Waltham, MA) containing 3 mL of viral transport medium.
Nasopharyngeal and anterior nasal swabs were used to sample both nostrils. Swab samples were vortexed vigorously for 10 seconds, aliquoted, and stored at –80°C prior to analysis.
Saliva was self-collected by participants via passive drool into a collection cup, then aliquoted and stored at –80°C prior to analysis.
Serum was collected using serum separator tubes, aliquoted, and stored either refrigerated or at –80°C until analysis by hemagglutination inhibition (HAI) assay.

**Aerosol Generation & Delivery**

<u>Inoculum preparation</u>

The $3{\cdot}6x10^5$ $TCID_{50}$/mL influenza A/Perth/16/2009 (H3N2) challenge stock, provided in a phosphate-buffered saline containing 25% sucrose (Meridian Life Sciences), was diluted prior to aerosolization for inoculation. The 1:100 dilution used with the FMAG was diluted in sterile injection water (Hospira Inc., Lake Forest, IL) and 10% Sweet-Ease natural sucrose solution consisting of 24% sucrose in a citric acid solution (International Biomedical, Austin, TX). The 1:100 dilution used with the medical nebulizer was diluted in Lactated Ringer's solution (ICU Medical, San Clemente, CA) and 9% phosphate-buffered saline solution containing 25% sucrose (manufactured by Naobios, France, on behalf of hVIVO, London, United Kingdom). The 1:10 dilutions used for both the FMAG and medical nebulizer were diluted in Lactated Ringer's solution. Diluted virus inoculum was transferred into syringes and stored on ice until administration. All inoculations were performed within three hours of thawing the stock virus.

<u>Inoculation using the FMAG</u>

Aerosolized virus was generated using a flow-focusing monodisperse aerosol generator (FMAG, TSI 1520) that produced particles 8-11 μm in size (Fig. S2). Participants were seated comfortably and wore a fit-test hood (3M) with a 1-inch barbed tube fitting attached to the hood's inlet. A three-foot corrugated hose connected the FMAG outlet to the barbed fitting on the hood. Airflow to the FMAG was provided by a compressed breathable air tank. A syringe (Henke-Ject, Seoul, Republic of Korea) containing at least 6 mL of the diluted virus inoculum was attached to the sample inlet port and the injection pump. The FMAG operated with a dilution air flow rate of 14-15 L/min, a focusing air pressure of 1.5 psi, a liquid concentration of 3.4%, a liquid flow rate of 6 mL/hr (0.1 mL/min), and a frequency of 80 kHz for the 1:100 virus inoculum dilution and 150 kHz for the 1:10 virus inoculum dilution. The FMAG was run for 20 minutes for the 1:100 virus inoculum dilutions and 10 minutes for the 1:10 virus inoculum dilutions. During this time, the participants were instructed to breathe normally. After this time, aerosol generation was stopped and participants removed the fit-test hoods.

<u>Inoculation using a medical nebulizer</u>

Aerosolized virus was generated using a medical jet-style nebulizer that produced smaller particles,<1-2 μm, in size (Fig. S2). Research participants were seated comfortably and wore a fit-test hood (3M) with a 0.75-inch barbed tube fitting attached to the hood's inlet. The nebulizer system consisted of an Aeromist Compact Compressor (Medline Industries, Northfield, IL) connected via plastic tubing to a Sidestream Plus nebulizer (Philips Respironics) The nebulizer contained 5 mL of diluted virus inoculum and was connected by a short length of plastic tubing to a barbed fitting on the hood. The nebulization system consumed liquid at a rate of ~0.1 ml/min and produced an aerosol flow rate of ~5 L/min. The compressor was turned on for 10 minutes during which the participant was instructed to breathe normally. Afterward, the participant then kept the fit-test hood on for an additional 10 minutes.

Estimation of exposure doses

After inoculations, replicate experiments were performed within the hood and without participants to measure the number of infectious viral units present in aerosols generated by the FMAG or the medical nebulizer. For the FMAG, these experiments used the same syringes containing the diluted virus inoculum that were used with the participants. Aerosols released within a fit-test hood were collected on a 37 mm diameter, 3.0 µm pore size PTFE filter (Millipore, Burlington, MA) connected to a vacuum pump with a flow rate of 5.0 L/min for 10 minutes. Virus was eluted from the PTFE filters by vortexing vigorously for 1 minute in 1 mL of universal viral transport medium (BD Biosciences, Franklin Lakes, NJ). For the medical nebulizer, a new Sidestream Plus nebulizer containing 5 mL of the diluted virus inoculum was used. The compressor was run for 10 minutes and aerosols generated within the fit-test hood were collected in phosphate buffered saline containing 0.5% bovine serum albumin and 0 ·2 M sucrose using a BioSpot-VIVAS (Aerosol Devices, Fort Collins, CO) running at an air flow rate of 8.0 L/min during nebulization and for an additional 10 minutes after. Infectious virus concentrations within these samples were measured by plaque assay. The estimated exposure dose was calculated according to Eq. 1,

$$D = C_{air} V_m t \quad (1)$$

where $D$ is the dose in PFU, $C_{air}$ is the concentration of virus in PFU/L air collected, $V_m$ is the participant's minute ventilation in L/min, and $t$ is the exposure duration in minutes. The concentration of virus in the air was determined from samples collected with the PTFE filter or BioSpot-VIVAS, and the participants' ventilatory volumes were measured using a spirometer (ndd Easy-on PC) while breathing normally with their noses clamped.

Determination of particle sizes generated during aerosolization

Particle size distributions generated during aerosolization of a buffered 2.5% sucrose solution were determined for both the FMAG and the medical nebulizer, with the same instrument settings as during participant inoculations, using an Aerodynamic Particle Sizer (TSI Inc. 3321). Fig. S2 shows example size distributions produced by both devices.

**Virus quantification**

Nasopharyngeal swab samples were analyzed using the diagnostic Xpert® Xpress SARS-CoV-2/Flu/RSV test on the GeneXpert Infinity platform (Cepheid, Sunnyvale, CA) at the Emory University Hospital microbiology laboratory**.** Infectious influenza virus concentrations were measured by plaque assay on Madin-Darby canine kidney (MDCK) cells as previously described[5]. Saliva samples with high viscosity were incubated with 0.05% dithiothreitol at 37°C for 10 minutes prior to plaque assays. Concentrations of influenza virus genome copies were measured by extracting nucleic acids from 160 µL samples using the NucleoMag RNA kit (Machery-Nagel, Dueren, Germany) on an epMotion 5075 (Eppendorf, Hamburg, Germany), followed by quantitative reverse-transcription polymerase chain reaction (qRT-PCR) as previously described *(5)*.

**Serum antibody measurement**

Hemagglutination Inhibition (HAI) assays were used to measure the levels of antibodies in serum that target the influenza A/Perth/16/2009 (H3N2) virus. Briefly, 25 µL of serially diluted receptor-destroying-enzyme- and heat-treated sera were mixed with 25 µL of virus solution containing 4 hemagglutination (HA) units for 15 minutes followed by the addition of 50 µL of 0.75% turkey red blood cells (Lampire Biological Laboratories, Pipersville, PA). After incubation at room temperature for 40 minutes, plates were scored for the presence of hemagglutination and the HAI titer was calculated as the last serum dilution that inhibited hemagglutination.

### SUPPLEMENTARY FIGURES AND TABLES

| Cohort | N | Mean Age (Range) | Female (N, %) | White (N, %) | Black (N, %) | Other Race (N, %) | Latinx (N, %) |
|---|---|---|---|---|---|---|---|
| **FMAG** | 6 | 30.8 (24-42) | 3 (50.0%) | 3 (50.0%) | 3 (50.0%) | 0 (0.0%) | 0 (0.0%) |
| **Medical Nebulizer** | 8 | 37.0 (21-49) | 7 (87.5%) | 4 (50.0%) | 3 (37.5%) | 1 (12.5%) | 0 (0.0%) |
| **All** | 14 | 34.4 (21-49) | 10 (71.4%) | 7 (50.0%) | 6 (42.9%) | 1 (7.1%) | 0 (0.0%) |

**Table S1 Demographic Table**

FMAG=flow-focusing monodisperse aerosol generator

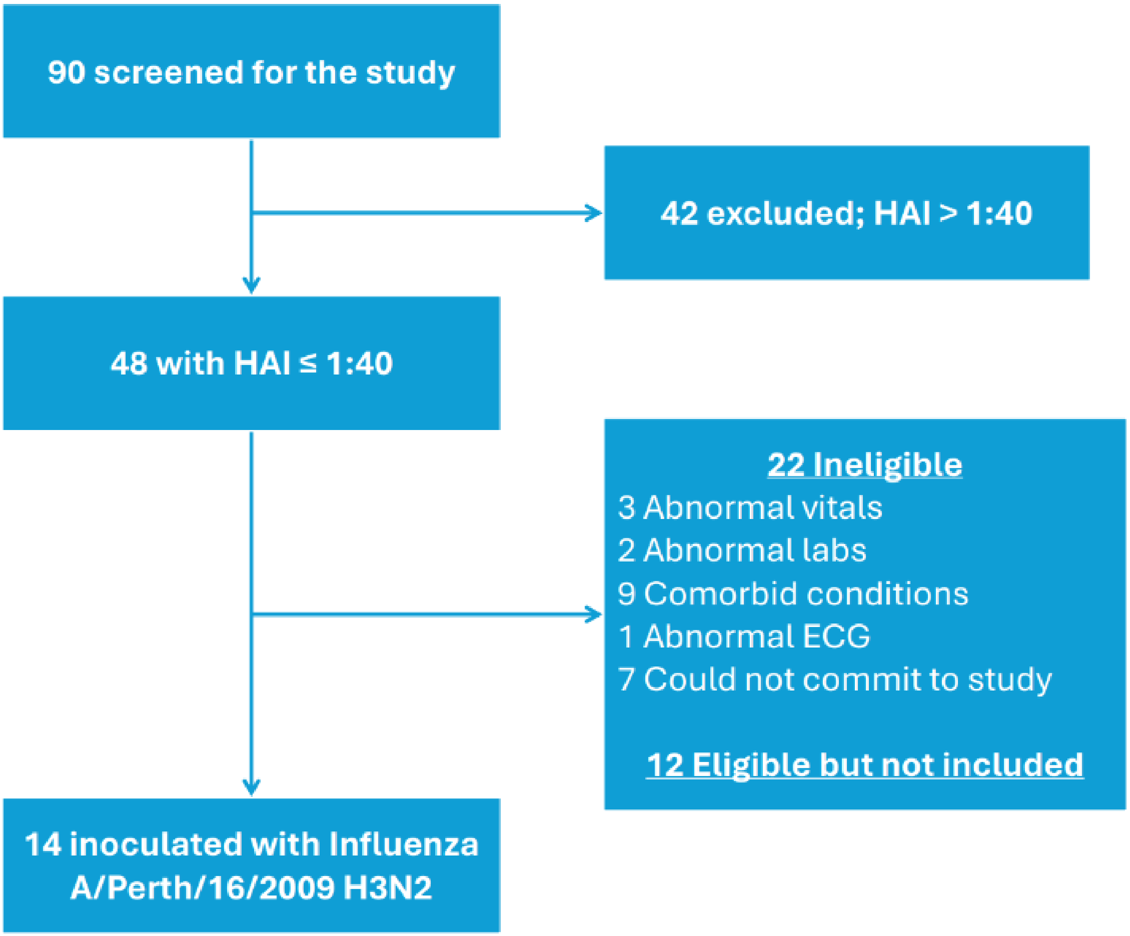


**Fig. S1 Consort diagram**

HAI: Hemagglutination Inhibition Assay; ECG: electrocardiogram.

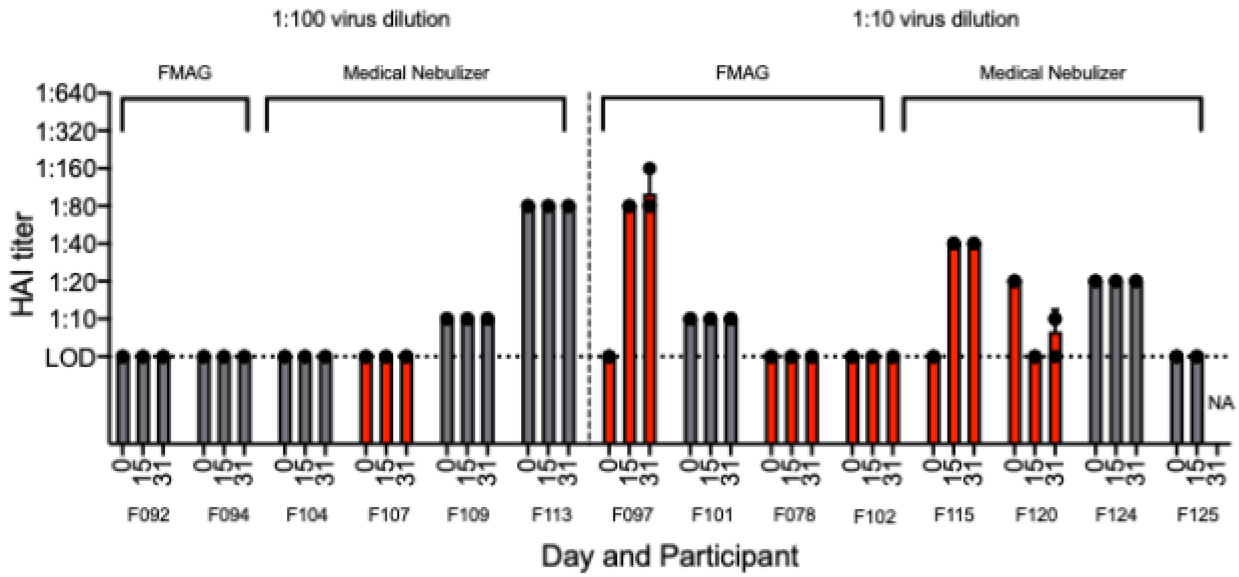


**Fig. S2 Serological responses to influenza A/Perth/16/2009 (H3N2)**

Sera collected from study participants on Days 0 (pre-inoculation), 15 (±3), and 31 (±3) were tested for antibodies capable of inhibiting hemagglutination of red blood cells in the presence of influenza A/Perth/16/2009 (H3N2) virus. Each sample was tested in triplicate. The geometric mean of the three replicates is plotted, with error bars representing the standard deviation. Red bars indicate participants with MMID (mild to moderate influenza disease). The dashed line represents the limit of detection (LOD). Participant F113 had an HAI titer of 1:80 on Day 0 despite a pre-study screening titer of 1:40. Additionally, participant F125 did not have a Day 31 visit and this sample was not available (NA) for testing. FMAG=flow-focusing monodisperse aerosol generator.

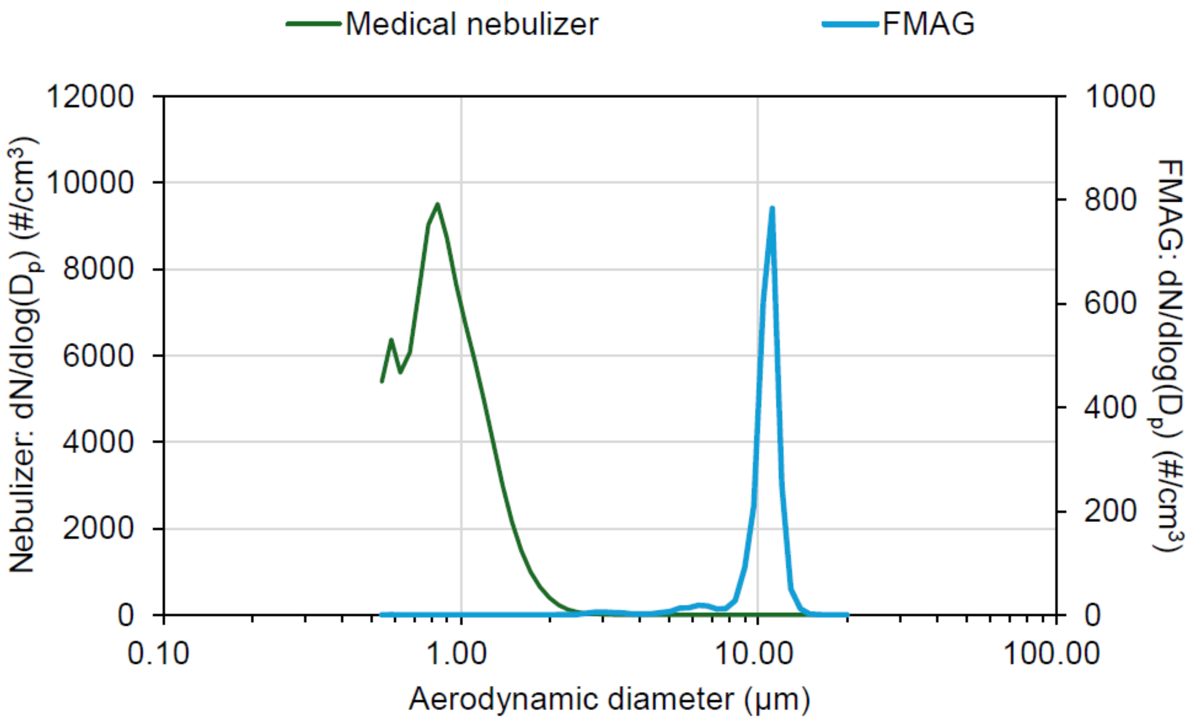


**Fig. S3 Particle size distributions of Medical Nebulizer and FMAG**

Example of particle size distributions produced by the medical nebulizer and FMAG (flow-focusing monodisperse aerosol generator) from a buffered 2.5% sucrose solution.
